# Cerebrospinal fluid proteomics reveals synaptic and immune dysregulation in treatment-resistant schizophrenia

**DOI:** 10.1101/2025.05.14.25327624

**Authors:** Laura E. Fischer, Michael G. Gottschalk, Lars U. G. Reitz, Joanna Moussiopoulou, Nikolaos Koutsouleris, Vladislav Yakimov, Emanuel Boudriot, Sophie Seeburger, Tengjia Jiang, Johanna Weiske, Agnese Petrera, Giuseppina Maccarone, CDP Working Group, Jaan-Olle Andressoo, Andrea Schmitt, Daniel Keeser, Alkomiet Hasan, Manfred Uhr, Peter Falkai, Sergi Papiol, Elias Wagner, Florian J. Raabe

## Abstract

**IMPORTANCE:** Treatment-resistant schizophrenia (TRS) affects around one-third of individuals with schizophrenia and is associated with substantial morbidity, mortality, and healthcare burden. Despite its impact, the pathophysiology of TRS remains poorly understood, and no validated biomarkers exist to predict or stratify treatment resistance and identify patients who may benefit from earlier second-line interventions.

**OBJECTIVE:** To identify cerebrospinal fluid protein signatures associated with treatment-resistant schizophrenia using high-throughput immune-based proteomic profiling, and to characterize their biological relevance and evaluate their predictive potential.

**DESIGN, SETTING AND PARTICIPANTS:** This cross-sectional study analyzed cerebrospinal fluid samples, collected between 2001 and 2024 during routine clinical practice in two centers, of 64 patients with treatment-resistant schizophrenia, and 84 sex-, age-, and center-matched non-treatment-resistant patients.

Proteomic profiling was performed using Olink Explore HT, a highly sensitive, multiplexed proximity extension assay capable of quantifying up to 5,416 proteins. Univariate differential expression analysis, Gene Set Enrichment Analysis, and Support Vector Machine modelling were conducted to identify treatment-resistant schizophrenia-associated proteins, explore underlying biological mechanisms, and uncover predictive biomarker signatures.

**MAIN OUTCOMES AND MEASURES:** Differentially expressed proteins, associated functional pathway enrichments, and classification performance of predictive models based on cerebrospinal fluid proteomics.

**RESULTS:** 1,839 protein assays passed detection threshold in cerebrospinal fluid proteomic analysis using Olink Explore HT. 98 proteins were differentially expressed in individuals with treatment-resistant schizophrenia and 95 displayed consistent fold changes across both study centers. Gene Set Enrichment Analysis revealed that upregulated proteins were linked to immune-related pathways, while downregulated proteins were predominantly enriched in synapse-related processes, including glutamatergic signaling.

Multivariate support vector machine modelling identified 64 proteins predictive of treatment-resistant schizophrenia, of which 63 overlapped with the differentially expressed proteins.

**CONCLUSIONS AND RELEVANCE:** Cerebrospinal fluid proteomic profiling indicates activation of immune-related processes and synaptic dysfunction in treatment-resistant schizophrenia, supporting the hypothesis that it represents a neurobiologically distinct subtype of the disorder. The identified candidate biomarker signature may support the development of liquid biopsy-based diagnostic tools and guide targeted treatment strategies.

**Key Points:** 

**Question:** Are there proteomic alterations in the cerebrospinal fluid (CSF) of patients with treatment-resistant schizophrenia (TRS)?

**Findings:** Unbiased CSF proteomics identified 98 proteins dysregulated in TRS, implicating pathways of neuroinflammation and synaptic dysfunction. Machine-Learning Validation pinpointed protein features with the strongest predictive value for TRS, of which 63 overlapped with the differentially expressed proteins.

**Meaning:** The intersection of univariate and multivariate results yields a prioritized panel of protein biomarkers with potential to distinguish TRS patients from those responsive to first-line antipsychotic therapy. The identified CSF-based proteomic signature reveals immune and synaptic alterations in TRS and may pave the way for early diagnostic tools and targeted interventions to alter its disease-course.

## Introduction

Treatment-resistant schizophrenia (TRS) affects around one-third of individuals with schizophrenia (SCZ)^1,2^ who suffer from persisting symptoms despite sequential trials of at least two different antipsychotics administered at appropriate doses and duration^2^. TRS is associated with functional impairment and elevated mortality, as well as a high socioeconomic burden^1,3–6^. Growing evidence suggest that TRS represents a distinct, yet internally also heterogenous, subtype of SCZ^7^. Prior studies identified TRS as a complex polygenic trait^8,9^ with dysregulated dopaminergic and glutamatergic signalling^10,11^, lipidomic alterations^12^, signs of immune system abnormalities^13^, and variations in brain circuits^10^.

Clozapine (CLZ) is the only approved medication for TRS^14^, but its application is often delayed by years because of rare safety concerns^15^ that have led to monitoring requirements^16^. Earlier consideration of second-line strategies such as clozapine could improve disease outcomes^17^, given the high frequency if TRS^18^. Therefore, timely identification of individuals at high risk for TRS via biomarkers is a critical unmet need.

Currently, most biomarker research in TRS often relies on hypothesis-driven approaches focusing on narrow sets of blood-based biomarker candidates^19–22^. This strategy risks to overlook informative unknow and/or novel biomarkers and is also challenged by the biological heterogeneity of TRS and the incomplete understanding of its underlying neurobiology. Considering the limitations of blood-based liquid biopsies in disorders of the central nervous system (CNS), cerebrospinal fluid (CSF) as the accessible fluid of the CNS could provide deeper insights into the pathophysiology of TRS and support biomarker discovery.

The German national schizophrenia guideline^23,24^ recommends a lumbar puncture as a standard differential diagnostic procedure for routine clinical practice in schizophrenia. The CSF obtained represents a unique resource for research. The advent of high-throughput proteomic technologies allows now the simultaneous quantification of thousands of proteins in small CSF volumes in the range of µLs.

To investigate TRS pathophysiology and to explore TRS biomarker candidates, we applied high-throughput proteomic technology to simultaneously quantify thousands of proteins in CSF from patients with SCZ. In this bi-centric study, we employed the Olink Explore HT platform, a high-throughput immunobased proximity extension assay combined with next-generation sequencing^25–27^, to measure the expression of 5,416 proteins in CSF samples from patients with TRS and non-TRS. Thereby, we aimed to 1) identify differentially expressed proteins (DEPs) associated with TRS, 2) advance the understanding of biological processes in TRS, and c) explore the potential of CSF protein signatures to predict TRS (**Figure 1**).

**Figure 1:**
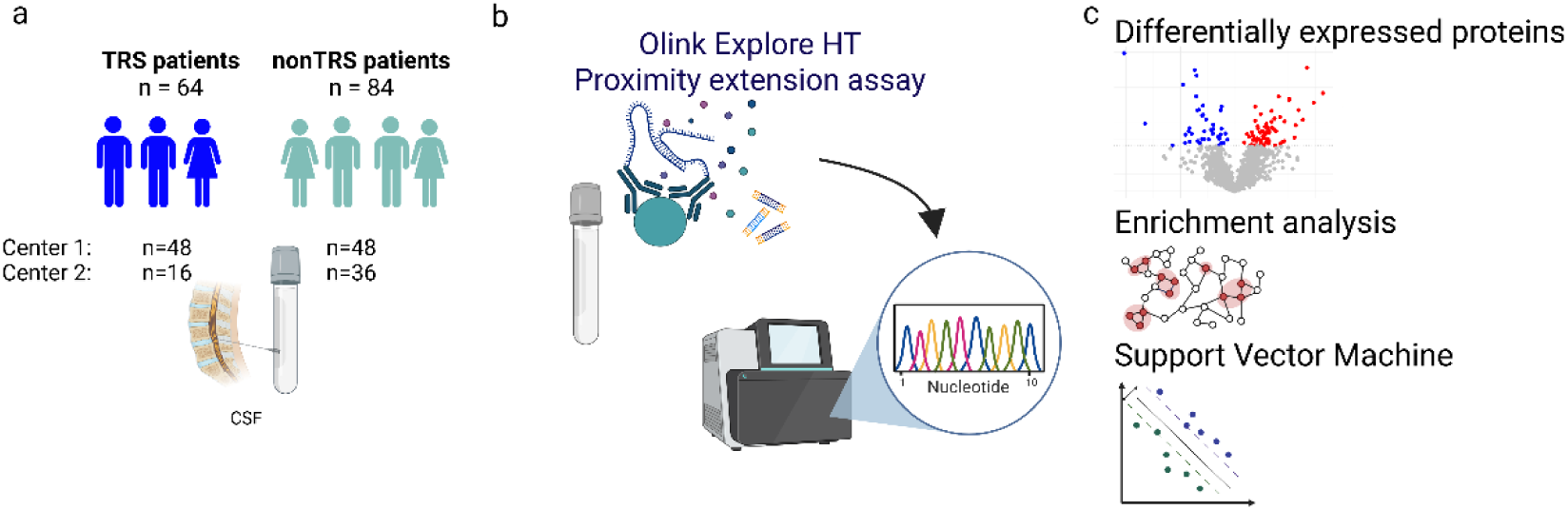
Schematic illustration outlining the proteomic investigation of cerebrospinal fluid in treatment-resistant schizophrenia. **a,** Cerebrospinal fluid (CSF) samples (n=148) were collected from patients with treatment-resistant schizophrenia (TRS) and non-treatment-resistant schizophrenia (non-TRS) across two centers. **b,** Proteomic profiling was conducted using high-throughput proximity extension assay technology with the Olink Explore HT assay. **c,** Overview of the analysis pipeline, including differential expression protein analysis, gene set enrichment analysis, and a machine learning workflow for identifying and characterizing candidate protein biomarkers for TRS. Figure was created with Biorender.com.

## Methods

### Study design and CSF collection

This cross-sectional study recruited mainly patients with SCZ (N=136) and some patients with schizophrenia spectrum disorders (N=12) from two independent study centers. CSF samples were collected during routine diagnostic lumbar puncture at the Max Planck Institute of Psychiatry, Munich, Germany (N=96) and the Department for Psychiatry and Psychotherapy, LMU University Hospital, Munich, Germany (N=52). Guided by the TRRIP (Treatment Response and Resistance in Psychosis) consensus criteria^2^ and in line with classification recommendations for real-world data^28^, participants were retrospectively classified as TRS or non-TRS based on study assessment and medical records **(eFigure S1A).** Moreover, CSF from patients with first-episode psychosis (FEP) was defined as collected CSF during the initial psychotic episode, while multi-episode psychosis (MEP) individuals had experienced more than one episode at the timepoint of lumbar puncture. All CSF donations were embedded in diagnostic lumbar punctures within the clinical routine care. For further details see **eMethods** in **Supplement 1**.

### Proteomics profiling

Proteomic profiling was performed with the Olink Explore HT platform (Olink Proteomics, Uppsala, Sweden), an immuno-based proximity extension assay combined with next-generation sequencing, which quantifies up to 5,440 proteins and enables the high-sensitivity detection of low-abundance proteins in small sample volumes^25–27^. Sample randomization was performed regarding TRS/non-TRS, center, sex, and age to minimize potential batch effects. Experiments were performed according to manufacturer’s protocol at the Olink certified laboratory of the Metabolomics and Proteomics Core, Helmholtz Zentrum Munich - German Research Center for Environmental Health, Neuherberg, Germany. Olink’s log2-transformed Normalized Protein Expression (NPX) values were used for relative quantification^26^. Only proteins that exceeded the limit of detection (LOD) in at least one sample as determined by the *OlinkAnalyze R package (v* 4.2.0), were included in the analysis (**eMethods** in **Supplement 1**).

### Statistical analysis and machine learning

For details see **eMethods** in **Supplements 1**, in shot: Differential expression analysis was conducted using *limma*^29,30^, with multiple testing correction via the *simpleM* approach to account for correlated features^31^. Annotations of the DEPs was performed using the *Open Targets Platform*^32^ and *Olink Insight Data Analysis Platform*. Gene Set Enrichment Analysis (GSEA) was performed based on the Gene ontologies “Biological processes”, “Molecular function”, and “Cellular components”^33,34^. Synaptic characterization of the DEPs was performed with *SynGO*^35^, the genetic enrichment of the DEPs was assessed using MAGMA (Multi-marker Analysis of GenoMic Annotation, version 1.09)^36^ with summary statistics from schizophrenia^37^ and treatment-resistant schizophrenia^38^ genome-wide association studies. To assess the predictive value of the CSF proteome, we used the machine learning toolkit NeuroMiner (version 1.3; https://github.com/neurominer-git/NeuroMiner_1.3) to train a L2-regularized support vector machine (SVM) classifier on the normalized NPX values above detection threshold. NPX values were adjusted for age and sex via partial correlation analysis. Univariate feature ranking using f-score was performed at increasing percentile thresholds to select most predictive features for model training. All analysis steps were wrapped into a repeated nested cross-validation design (10×10 outer folds; 5×10 inner folds) to avoid information leakage. Permutation testing with label shuffling (1000 permutations) was employed to assess model significance, determined at α = 0.05, as described previously^39^.

## Results

### Cohort characteristics and assay detectability

The cohort included CSF from 148 patients, of whom 64 were characterized as TRS and 84 as non-TRS (**Figure 1A**). Sex distribution and age (mean 37 years) were not significantly different between TRS and non-TRS (**Table 1**). TRS patients had significantly more hospitalizations (p=.001; **Table 1**) and a higher BMI (p=.0143; **Table 1**) compared to non-TRS. Within the TRS group, 30 patients had CLZ treatment prior to or at the time of lumbar puncture, and seven patients were in their first-episode (**Table 1** and **eFigure S1B**). The total cohort included 34 individuals with FEP and 41 with MEP who were both treated with antipsychotic agents at the timepoint of study inclusion. For further details, see **Table 1** and **TableS1**.

**Table 1:** Baseline characteristics of patients with treatment-resistant schizophrenia (TRS) and non-treatment resistant schizophrenia (non-TRS) FEP first-episode patients. CPZ chlorpromazine. CSF cerebrospinal fluid. Ig G Immunglobuline G. ^a^n = 58; ^b^n = 69; ^c^n = 69; ^d^n = 78; ^e^n = 62; ^f^n = 82; ^g^n = 54; ^h^n = 61; ^i^n = 59; ^j^n = 67; ^k^n = 50; ^l^n = 63; ^m^n = 61; ^n^n = 82; °n = 62; ^p^n = 82; ^q^n = 61; ^r^n = 83; ^s^n = 61; ^t^n = 83; ^u^n = 60; ^v^n = 83. *Two-sided Fisher’s exact test was used for contingency tables or two-sided Mann–Whitney U test for continuous values.

| **Clinical characteristics** | **n** | **TRS** | **non-TRS** | **p-value<sup>TRS vs non-TRS</sup>** |
| --- | --- | --- | --- | --- |
| Cohort size (n) | 148 | 64 | 84 |  |
| Age (range), years | 148 | 38.0 (18, 73) | 34.5 (18, 70) | U=2985.50, p=.2501 |
| Sex (F/M) | 148 | 32/32 | 34/50 | Chi2=0.98, p=.3232 |
| CPZ aequivalent in mg (SD) | 131 | 820 (658) | 593 (507) | U=2985.5, p=.250 |
| Number of hospitalizations (n) | 127 | 3.0 (0.0, 21.0)<sup>a</sup> | 1.0 (0.0, 5.0)<sup>b</sup> | **U=3087.00, p=.0000** |
| Medicated FEP (Yes/No) | 138 | 7/53<sup>c</sup> | 33/45<sup>d</sup> | **Chi2=14.02, p=.0002** |
| Clozapine current (Yes/No) | 144 | 29/33<sup>e</sup> | 0<sup>f</sup> | **Chi2=41.23, p=.0000** |
| Clozapine in history (Yes/No) | 115 | 26/28<sup>g</sup> | 0<sup>h</sup> | **Chi2=31.95, p=.0000** |
| BMI in kg/m<sup>2</sup> (SD) | 126 | 26.6 (4.7)<sup>i</sup> | 24.6 (4.3)<sup>j</sup> | **U=2478.00, p=.0143** |
| Current Smoking (Yes/No) | 113 | 21/29<sup>k</sup> | 31/32<sup>l</sup> | Chi2=0.33, p=.5664 |
| **CSF basic assessment** |  |  |  |  |
| CSF cell count (range) | 143 | 2.0 (0.0, 16.0)<sup>m</sup> | 1.0 (0.0, 11.0)<sup>n</sup> | U=2923.50, p=0.0775 |
| Erythrocyte count <500 (%) | 144 | 100<sup>o</sup> | 100<sup>p</sup> |  |
| Albumin ratio (range) | 144 | 6.27 (2.39, 15.9)<sup>q</sup> | 5.68 (2.51, 14.06)<sup>r</sup> | U=2896.00, p=0.1411 |
| IgG ratio (range) | 144 | 3.08 (0.27, 11.42)<sup>s</sup> | 2.69 (0.98, 7.59)<sup>t</sup> | U=2763.00, p=0.3503 |
| Oligoclonal CSF IgG bands (Yes/No) | 143 | 6/58<sup>u</sup> | 5/79<sup>v</sup> | t=0.78, p=0.4350 |

Nearly all 5,416 assayed proteins in CSF using the Olink Explore HT platform (**Figure 1B**) passed quality control, only ApoE was excluded due to elevated negative-control counts. Based on LOD determination according to manufacturer’s recommendations, 1,839 proteins reached the LOD threshold and were included in downstream analysis (**eFigure S2A**).

### CSF-derived proteomic alterations indicate immune and synaptic dysregulation in TRS

To investigate CSF proteomic alterations in TRS, we performed differential expression analysis using linear model t-test (Limma) and identified 98 significant DEPs (**Figure 2A**, **Table S2**). We assessed robustness by comparing fold-change directions across both independent centers and found 95 of the 98 DEPs to be consistently dysregulated (**Figure 2B**). Notably, merely a handful of DEPs, including C4B, IL18, GRIK2, KCNC4, and NLGN1 had been previously linked to SCZ or TRS (**Table S3**)^32^, while the majority of DEPs was newly discovered. Due to the highly correlated proteome (**eFigure S2B**), multiple-testing correction was performed via the SimpleM method^31^. Thereafter, only fatty acid binding protein 4 (FABP4, upregulated), Ependymin Related 1 (EPDR1) and Arylsulfatase B (ARSB) (both downregulated) remained significant. This underscores the challenge of simultaneously assaying thousands of proteins, an achievement made possible by advanced high-throughput proteomic platforms, in relatively small cohorts. Moreover, it has to be considered that overly conservative thresholds can obscure true signals ^40^.

**Figure 2:**
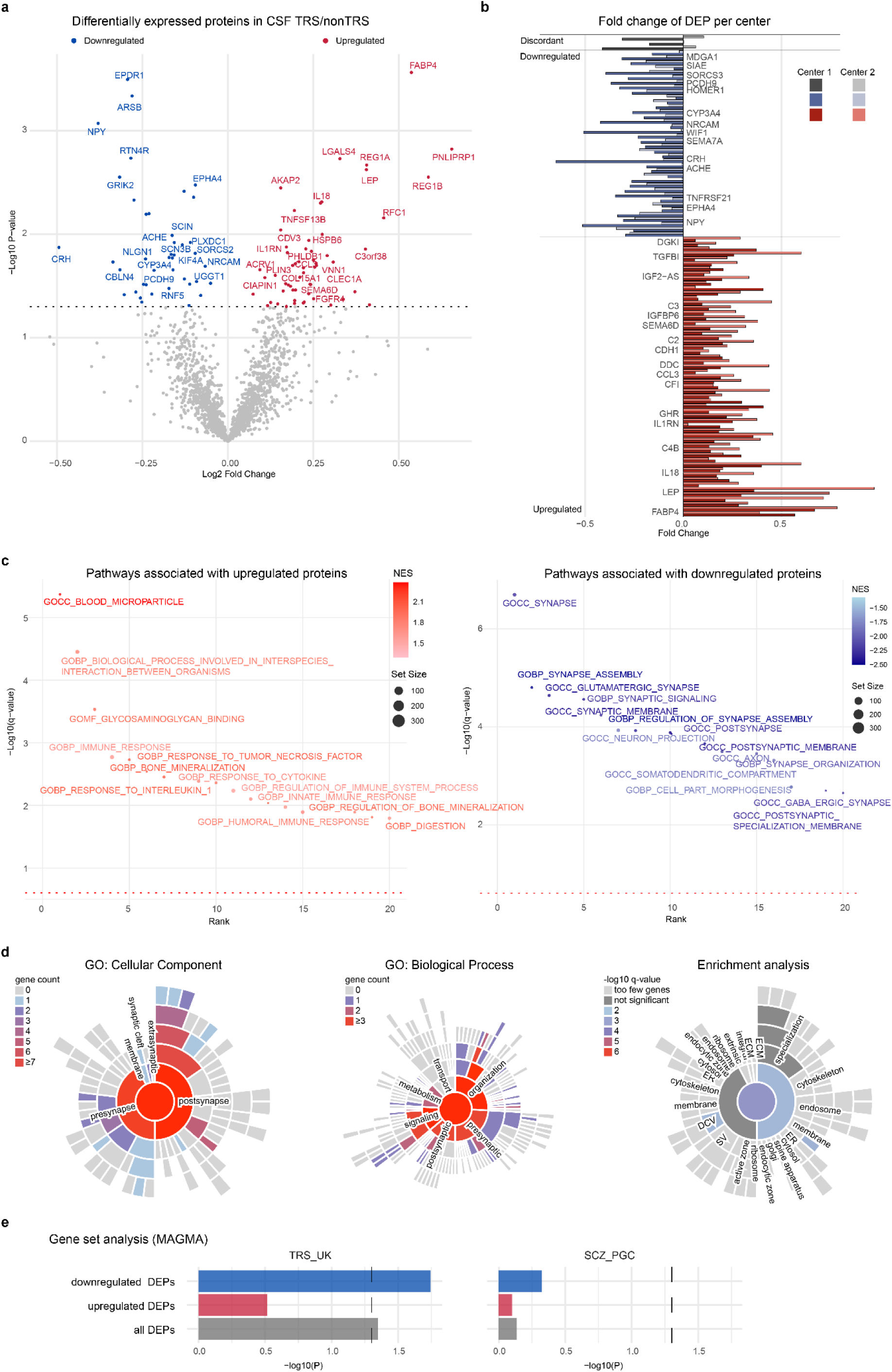
Differentially Expressed Proteins in treatment resistant schizophrenia are linked to immune- and synapse-related dysregulations. **a,** Volcano plot displaying the result of differential expression analysis in CSF, conducted using a two-sided linear model t-test (Limma) adjusted for sex, age, and center. Differentially expressed proteins (DEP) with significant upregulation (red) and downregulation (blue) in treatment resistant schizophrenia (TRS) exceeding a p-value threshold of 0.05 are color-coded, while non-significant proteins are shown in grey. **b,** Bar chart of the log2 transformed ratio of the mean expression of NPX values of the significant Explore HT assays between TRS patients and non-TRS patients (logFC) of the DEP, displayed for both centers individually. The y-axis orders protein assays by p-value magnitude, with smaller p-values positioned further up, separately for discordant (opposing fold-change-directions, n=3), consistently upregulated (n=57) and consistently downregulated (n=38) proteins across the two study centers. **c,** Scatter plot illustrating Gene Ontology (GO) terms of Biological Processes (BP) Cellular Components (CC) and Molecular Functions (MF) pathways associated with upregulated (red) or downregulated proteins (blue) in treatment-resistant schizophrenia (TRS). The enrichment analysis was based on fold changes and p-values obtained from Limma analysis of 1839 proteins, examining 2,288 GO gene sets. The red line represents a q-value threshold of 0.25. Color gradient indicates the normalized enrichment score (NES), and circle size corresponds to the number of proteins associated with the respective GO term. Q-value is used as Rank criterium. **d,** Sunburst plot from SynGO illustrating cellular component (left, n=21) and biological process (centered, n=17) annotation of DEP in SynGO ontology. Colors indicate gene count per term. SynGO-enrichment analysis (right) identified four cellular components (CC) and one biological process (BP) to be significantly enriched after FDR-correction (testing terms with at least three matching input genes). Colors indicate FDR value. **e,** MAGMA enrichment analysis of DEP coding genes in TRS and SCZ GWAS. Results show enrichment of DEP gene sets in TRS but not in SCZ.

As individual genes do not fully reflect the effects of disturbed biological processes, we performed GSEA {Subramanian, 2005 #426} on the full proteome above LOD across GO Molecular Function, Biological Process, and Cellular Component categories^33,34^ to reveal underlying biological information in TRS.

Upregulated proteins were strongly enriched in immune-related pathways, including innate and adaptive immunity, immune regulation, and inflammation. Moreover, glycosaminoglycan binding and extracellular matrix organization were altered in SCZ (**Figure 2C**, left panel, and **Table S2)**. In contrast, downregulated proteins mapped significantly to synaptic processes, such as synapse assembly, and glutamatergic transmission (**Figure 2C**, right panel, and **Table S2**). Deeper characterization of the synaptic proteins revealed alterations across multiple synaptic processes and functions, with pronounced effects at the postsynaptic level (**Figure 2D**). Next, we investigated the genetic background of the identified DEPs with MAGMA and revealed a significant enrichment of the DEP gene set, particularly the downregulated set, in the genetic architecture of TRS but not in SCZ, highlighting the genetically driven differences in TRS (**Figure 2E** and **Table S4**).

### Proteomic subgroup profiling indicates specificity of the TRS-DEPs

To assess whether our TRS-DEP signature captures treatment resistance rather than general disease chronicity, we derived a chronicity-related proteomic profile by comparing CSF from treated FEP to that of treated MEP patients. We then compared this chronicity signature with the TRS-specific DEPs to determine which proteins uniquely characterize treatment resistance versus those reflecting prolonged illness and medication exposure. We identified 70 upregulated and 164 downregulated proteins in FEP compared to MEP patients (**eFigure S3A**). Remarkably, only six proteins overlapped with TRS-DEPs (**eFigure S3B**), indicating that proteomic alterations associated with disease chronicity are largely distinct from those linked to treatment resistance.

Acknowledging both CLZ’s potential impact on the CSF proteome and its typical use in individuals with higher SCZ polygenic risk scores^41^, and given that half of the people with TRS in our study received CLZ at the time of CSF collection, we also compared the proteomic profiles of individuals receiving CLZ at the time point of lumbar puncture versus those patients not receiving CLZ. This analysis revealed 170 upregulated and 127 downregulated proteins in CLZ-treated individuals (**eFigure S3C**). Notably, 44 DEPs overlapped between the CLZ-DEPs and TRS-DEPs signatures (**eFigure S3D**), reflecting the clinical intersection between CLZ treatment and treatment resistance.

To further account for potential medication effects on the CSF proteome, we investigated whether total antipsychotic dose, quantified as CPZ equivalents, influenced the magnitude of NPX expression of the DEPs. Antipsychotics, particularly at higher doses, may exert widespread neurochemical effects that could confound biological signals associated with treatment resistance. To assess this, we correlated CPZ equivalent dosages with NPX values of the 98 identified DEPs. Notably, only 5 out of the 98 proteins showed a significant correlation with CPZ equivalent dosages (**Table S5**). Furthermore, only 3 out of 98 proteins showed a significant NPX correlation with CLZ daily dose (**Table S5**).

### Performance of CSF proteins to predict TRS

In order to evaluate the potential of the CSF proteome for personalized medicine approaches, we applied a SVM model, revealing significant predictive potential of the CSF proteome to identify patients with TRS with a balance between the True Positive Rate (TPR) and False Positive Rate (FPR; P<.001; **Figure 3A**). However, the balanced accuracy (BAC) was 63.6% showing a low to moderate ability to balance the classification of both classes (AUC =0.64, 95%CI:0.55-0.73; **Figure 3B**). Out of the 64 most predictive features of the SVM model (**Figure 3C**, **Table S6**), 63 proteins overlapped with the 98 DEPs identified in the univariate DEP analysis (**Figure 3D**), highlighting these as candidate biomarkers for identifying TRS among patients with SCZ.

**Figure 3:**
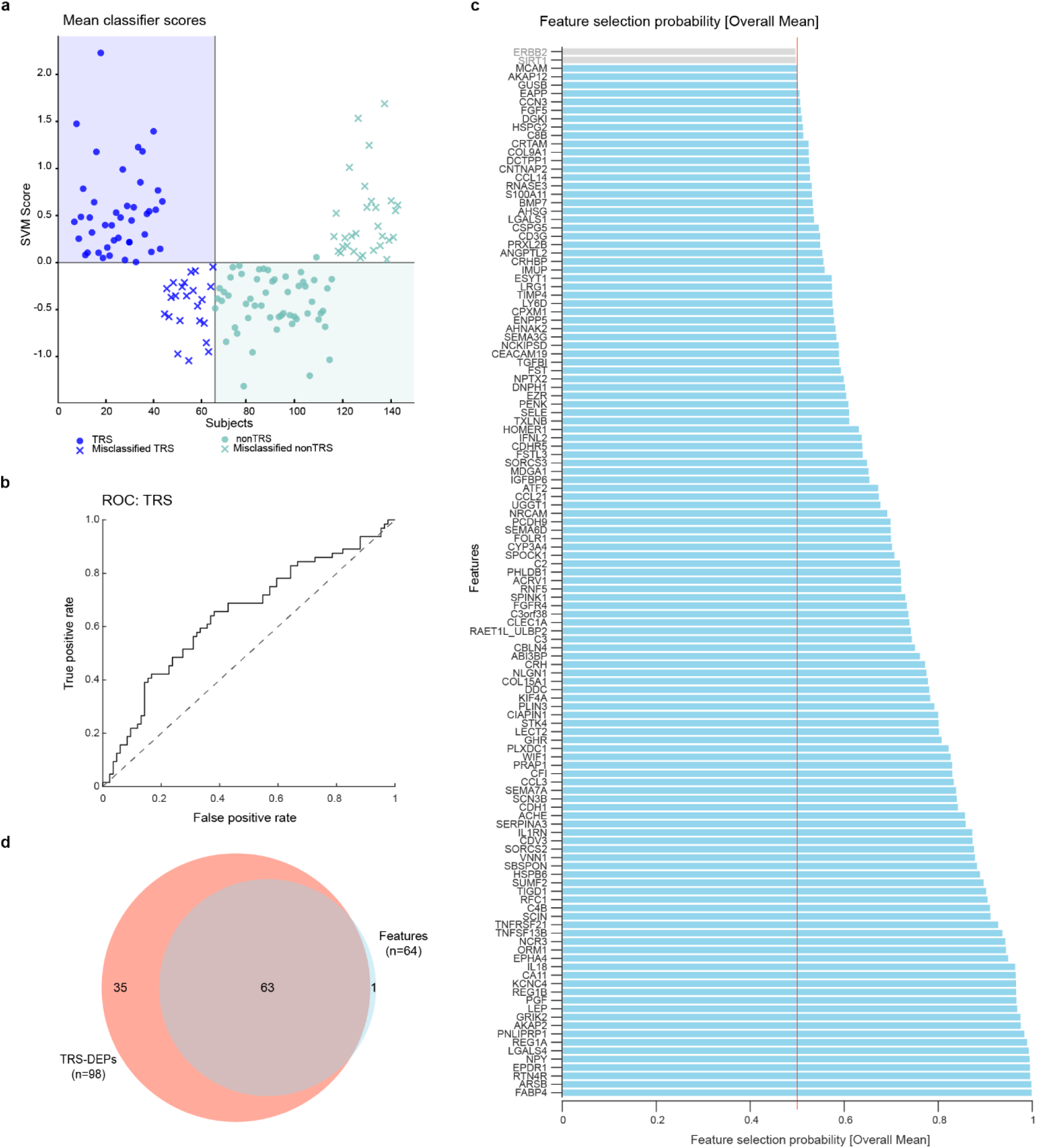
Predictive performance of cerebrospinal fluid (CSF)-derived proteins to identify patients with treatment-resistant schizophrenia. **a,** Support Vector Machine (SVM) decision scores highlighting the model’s ability to differentiate treatment-resistant schizophrenia (TRS, blue) from non-TRS patients (green). Dots illustrating correctly classified individuals and crosses visualize false-negative and false-positive TRS-classified individuals with schizophrenia. **b,** The receiver operating characteristic (ROC) curves illustrating the SVM model performance to separate TRS patients (n = 64) and non-TRS patients (n = 84) based on CSF proteome (AUC = 0.64, 95% CI: 0.55–0.73). Of all people with TRS, 64.1% were correctly classified predicted as TRS. For people with non-TRS, 63.1% were correctly identified. **c,** Bar plot chart illustrating the features (proteins) of the SVM analysis that have a sign-based consistency p-value < 0.05 after FDR correction across all SVM models, emphasizing the most robust contributors to the model’s predictive performance. **d,** Venn diagram showing the overlap of the 98 TRS-DEP identified by univariate two-sided linear model t-test (Limma) analysis with the top 64 explanatory significant features of the SVM model.

## Discussion

In this proteomic investigation, we profiled the CSF from individuals with TRS and matched non-TRS samples using a state-of-the-art proteome analysis platform, that can robustly quantify even low-abundance proteins.

Out of 1,839 measurable proteins, we revealed 98 proteins altered in TRS. Although only three DEPs remained significant after multiple-testing correction, 95 DEPs showed a consistent fold change direction across both centers, indicating a robustness of the identified signature. Characterizing the identified TRS-associated proteins, only a few had previously been linked to TRS, underscoring the potential of unbiased methods to uncover novel biomarker candidates.

In line with hypothesized neurobiological etiologies of TRS (Potkin, Kane et al. 2020), our GSEA of the CSF proteome uncovered a robust enrichment of immune-related processes in TRS. Notably we see high enrichment for the “Immune Response” gene set, including chemokines (CCL3, CCL8, CCL21), complement factors (C3, CFB), and interleukins (IL-1B, IL-18, IL-1RN). We also identified enrichment of the “Response to Tumor Necrosis Factor” pathway, driven by elevated expression of TNF-responsive genes such as SELE, TNFSF13B and ADAMTS13, pointing to pro-inflammatory gene expression (via NF-κB) and induction of apoptosis, accompanied by IL-1 pathway regulators (IL1RN, IL1R2). These findings underscore pervasive innate immune and cytokine signaling activity in TRS CSF, corroborating reports of peripheral Th17/IL-23 axis dysregulation^13,42^ and immune dysregulation in TRS^42^. Of note, several complement cascade components (C3, C4B, C2, and the regulatory factor CFI) were among the up-regulated proteins, which could also reflect dysregulated microglial or complement-mediated synaptic pruning^43^.

Proteins involved in the assembly, plasticity, and function of synapses – a core processes disrupted in SCZ^44,45^ – were markedly downregulated in TRS. Key neuronal adhesion and synaptic scaffolding proteins, such as NLGN1, NRCAM, HOMER1, GRIK2, and CBLN4, showed reduced levels, in line with previous descriptions of excitatory/inhibitory imbalance and impaired synaptic scaffolding in TRS^46^. In addition, several postsynaptic proteins, such as CDH2, EPHA4, GRIK2, GRIK5, SNAP25, SORCS3, and SPOCK1, were downregulated in the CSF of patients with TRS, in agreement with postmortem evidence of diminished postsynaptic density in prefrontal cortical regions of SCZ patients^47^. Interestingly, several dysregulated proteins were linked to glutamatergic signaling, which has been specifically implicated in the pathophysiology of TRS^48,49^.Taken together, the various findings of our study could point to an immune–synaptic pathology in TRS: chronic inflammatory signaling in addition to elevated complement activity may drive exacerbated synaptic loss in critical cortical circuits, perpetuating treatment refractoriness. This integrated model aligns with genetic studies showing that variation at the at the MHC locus, specifically in the two human C4 genes, modulates risk for SCZ and that neuronal C4 protein localizes to dendrites, axons and synapses where it promotes microglia-mediated synapse elimination during adolescence^50^. Our data support the notion that dopamine-targeted treatments alone might be insufficient to restore synaptic integrity in TRS^44^.

Strikingly, our genetic enrichment analysis revealed a significant association between TRS-DEPs and the genetic risk for TRS^38^, but not with the broader genetic architecture of SCZ. This genetic finding not only retrospectively supports the validity of our real-world TRS classification but also reinforces the hypothesis that TRS represents a neurobiologically distinct subtype of schizophrenia.

Importantly, by comparing the CSF proteomes of FEP and MEP, we were able to show that the TRS-associated proteomic changes are largely distinct from a protein signature reflecting disease chronicity. The very limited overlap suggests that chronicity-related proteomic changes in SCZ are largely distinct from those associated with treatment resistance and highlights the specificity of the TRS proteomic signal. Considering that around half of TRS patients received CLZ at the time point of lumbar puncture, we observed a substantial overlap between the TRS-associated proteomic alterations and the proteome signature distinguishing CLZ-treated from non-CLZ individuals. Given the challenge in disentangling disease-driven from medication-induced proteomic alterations - and the fact that individuals receiving CLZ suffer from a higher symptom burden^51^ - we cannot rule out that a subset of the observed TRS-associated proteomic alterations may be influenced by CLZ treatment which has broad biological effects^52–55^. However, the expression level of only 5% or fewer DEPs correlated with chlorpromazine (CPZ) or CLZ dosages, suggesting that the majority of the TRS signature is not a direct effect of antipsychotic exposure. Dissecting medication-vs. disease-driven proteomic changes will require future longitudinal and mechanistic studies.

Complementing univariate DE analysis, multivariate SVM analysis provided additional insights into the robustness of the identified DEPs. Although the classification performance exceeded chance levels, the model performance remains below the threshold required for routine clinical decision making, underscoring the need for multimodal biomarker integration to pave the way towards clinical utility. In sum, PEA targeted proteomics technology combined with next-generation sequencing on CSF samples from patients with TRS offers a direct window into CNS pathophysiology. Our data analysis collectively underscores the multifaceted nature of TRS as a distinct subtype of general SCZ and highlights both immune dysregulation and synaptic impairments as important processes in its pathology.

### Limitations

Key limitations include a modest sample size, an inherent challenge in CSF research, high costs associated with modern proteomic techniques, and the retrospective design without prospective TRS criteria (with potential risk of misclassifying some TRS patients within the non-TRS group). Patient classification relied on TRRIP working-group definitions emphasizing pharmacological non-response. Current antipsychotic treatments are primarily effective for alleviating positive symptoms, but many patients continue to experience persistent negative symptoms and cognitive impairments. To identify biomarkers associated with the development of cognitive impairment in a specific subgroup, an alternative patient classification would be required. Additionally, although dosage correlations were minimal, the confounding effects of CLZ and other antipsychotics cannot be fully excluded and underline the need of longitudinal studies.

Unlike previous peripheral blood-based or hypothesis-driven approaches, our unbiased, high-throughput CSF proteomics investigation enables the identification of novel TRS-associated proteins that were consistently dysregulated across both centers. Moreover, by combining univariate and multivariate SVM analyses, we enhance the robustness of our findings and capture not only individual protein changes but could also reveal insights of the multifaceted biology of TRS. Importantly, our findings of immune and synaptic disruptions, particularly involving glutamatergic signaling, underscore the need for mechanistic studies to better understand the biology underlying TRS.

Finally, our promising findings require validation in independent and longitudinal cohorts. Integrating data from complementary proteomic platforms (e.g., mass spectrometry) and other -omics fields (e.g., metabolomics, genomics) may further elucidate TRS pathophysiology and support the development of (multiparametric) tools for early TRS prediction, with the ultimate goal of translating the CSF-based signatures into scalable blood-based applications.

## Conclusion

This study advances our understanding of TRS by implicating convergent immune and synaptic pathways in its pathophysiology, warranting follow-up mechanistic investigations. Additionally, this work identified a panel of CSF-derived biomarker candidates that may form the basis of an early-detection signature for TRS, with the potential to guide timely detection of at-risk individuals and to enable targeted personalized interventions in order to improve long-term outcomes in TRS.

## Data Availability

All data is available in the supplementary data. Raw data and CSF are available upon reasonable request that could require MTA/DTA-agreement with the authors. Please note that certain subsets of patient-derived data are subject to sharing restrictions due to limited donor consent. Moreover, additional ethics approval could be required dependent on the type of request.

## Code availability

All software packages used in this study are publicly available.

## Acknowledgments

The authors thank all participants for their contribution. Special thanks to Christian and Tamara Namendorf, Max Planck Institute of Psychiatry, Munich, Germany, for their help with sample preparation and to all members of the Core Unit “Analytics and Mass Spectrometry” at the Max Planck Institute of Psychiatry, Munich, Germany, for their efforts in CSF sample biobanking. Thanks also to Monika Rex-Haffner for support with organizational matters. We would also like to thank all colleagues at the Department of Psychiatry at the LMU Munich, Germany, and the Max Planck Institute of Psychiatry, Munich, Germany, for their efforts in conducting lumbar punctures and in biobanking CSF.

Given the—albeit still limited—data on preferred terminology, and to acknowledge both the protected status of individuals with severe illnesses and the long history of stigma experienced by those with psychiatric disorders, we will use the terms ‘patients with SCZ’, ‘individuals with SCZ’, and ‘people with SCZ’ synonymously throughout this document.

## Author contributions

Conceptualization: LEF, MU, PF, SP, EW, FJR; Data curation: LEF, NK, SP, FJR; Formal analysis: LEF, NK, TJ, SP, FJR; Funding acquisition: LEF, AS, PF, EW, FJR; Investigation: LEF, MGG, LUGR, JM, VY, EB, SS, AP, CDP, DK, MU, SP, EW, FJR; Methodology: LEF, NK, MU, PF, SP, FJR; Project administration: LEF, SP, EW, FJR; Resources: JM, NK, AS, AH, MU, PF, EW, FJR; Software: LEF, NK, TJ, JW; Supervision: FJR; Visualization: LEF, NK, FJR; Writing – original draft: LEF, SP, FJR; Writing – review & editing: All authors.

## CDP Working Group

Stephanie Behrens, Emanuel Boudriot, Man-Hsin Chang, Valéria de Almeida, Sylvia de Jonge, Fanny Dengl, Peter Falkai, Laura E. Fischer, Nadja Gabellini, Vanessa Gabriel, Sabrina Galinski, Thomas Geyer, Katharina Hanken, Alkomiet Hasan, Genc Hasanaj, Alexandra Hisch, Georgios Ioannou, Iris Jäger, Marcel Kallweit, Temmuz Karali, Susanne Karch, Berkhan Karslı, Daniel Keeser, Christoph Kern, Nicole L. Klimas, Maxim Korman, Nikolaos Koutsouleris, Lenka Krčmář, Constanze Lobinger, Isabel Lutz, Verena Meisinger, Julian Melcher, Matin Mortazavi, Joanna Moussiopoulou, Karin Neumeier, Frank Padberg, Boris Papazov, Irina Papazova, Sergi Papiol, Pauline Pingen, Oliver Pogarell, Siegfried G. Priglinger, Florian J. Raabe, Lukas Röll, Moritz J. Rossner, Philipp Sämann, Michelle Schamberger, Andrea Schmitt, Susanne Schmölz, Eva C. Schulte, Enrico Schulz, Benedikt Schworm, Elias Wagner, Sven Wichert, Vladislav Yakimov, Peter Zill.

## Supplementary information

Supplement 1

- Legends supplementary tables: TableS1, TableS2, TableS3, TableS4, TableS5

- separate Excel files
- eFigures 1–3
- eMethods

---

**eFigure S1:**
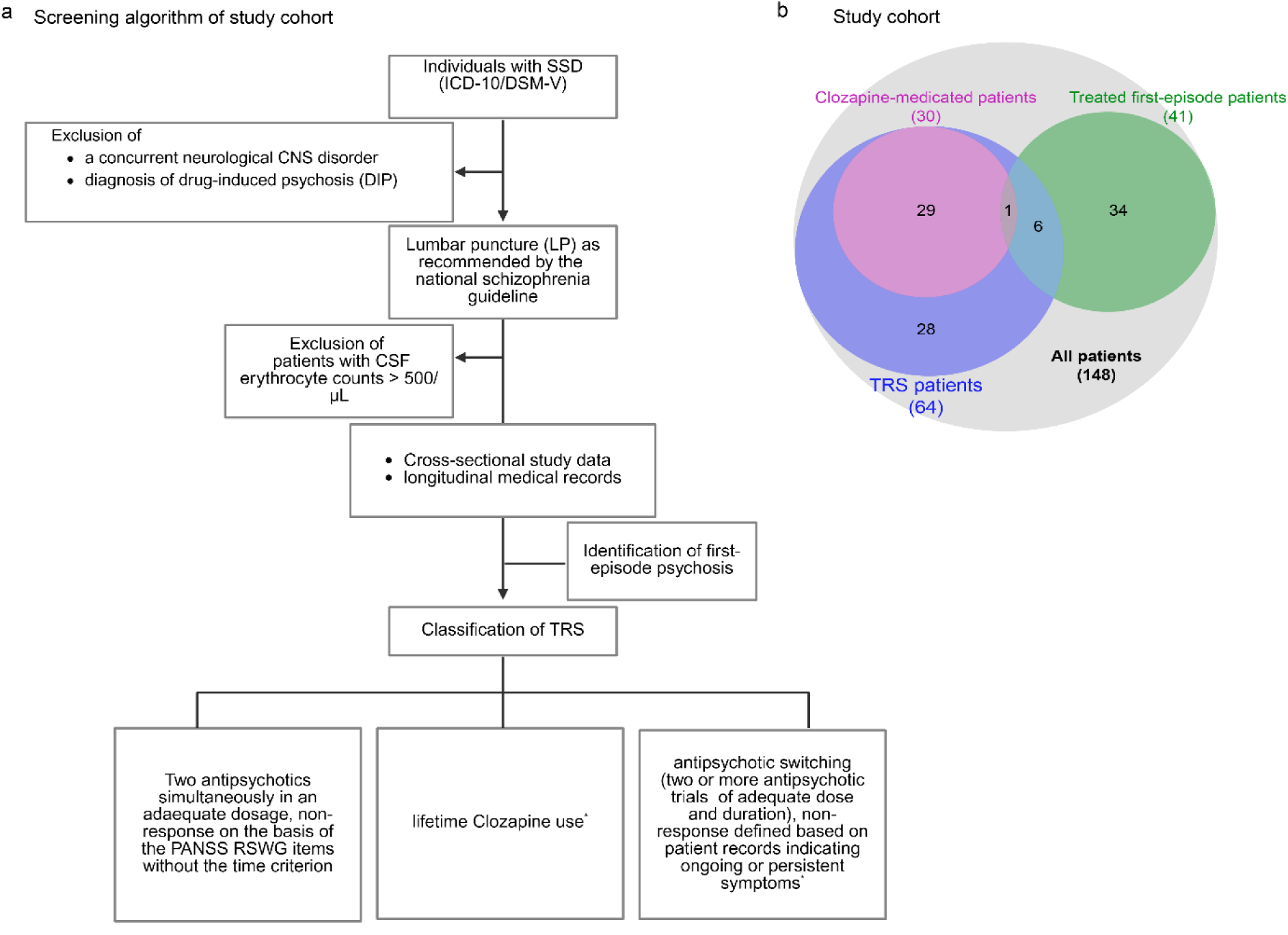
Flow chart of cohort design and schematic patients’ subgroup composition. **a,** Flowchart illustrating the screening algorithm used to define the final study cohort and description of TRS classification: 1. The use of two antipsychotic medications in an adequate dosage concurrently. Non-Remission was evaluated on the basis of the “Andreasen criteria” without the time criterion 2. Receiving CLZ treatment at any point in their lifetime or at LP 3. Antipsychotic therapy for ≥12 weeks, including at least six weeks with two different agents either concurrently or sequentially in an adequate dosage. Non-remission was determined by two trained psychiatrists through a comprehensive review of patient records. With regard to antipsychotic dosage, an equivalent dose ≥ 600mg of chlorpromazine equivalents per day was considered as a minimum requirement for treatment resistance based on established conversion criteria^56–60^. Classifications marked with an asterisk (*) align with recommendations for real-world data ^28^. **b,** Venn diagram illustrating the cohort composition and highlighting the overlap between TRS patients, patients receiving clozapine, and medicated first-episode psychosis (FEP) patients at the time point of lumbar puncture to collect the analyzed CSF. FEP were identified as those who underwent lumbar puncture during their initial psychotic episode.

**eFigure S2:**
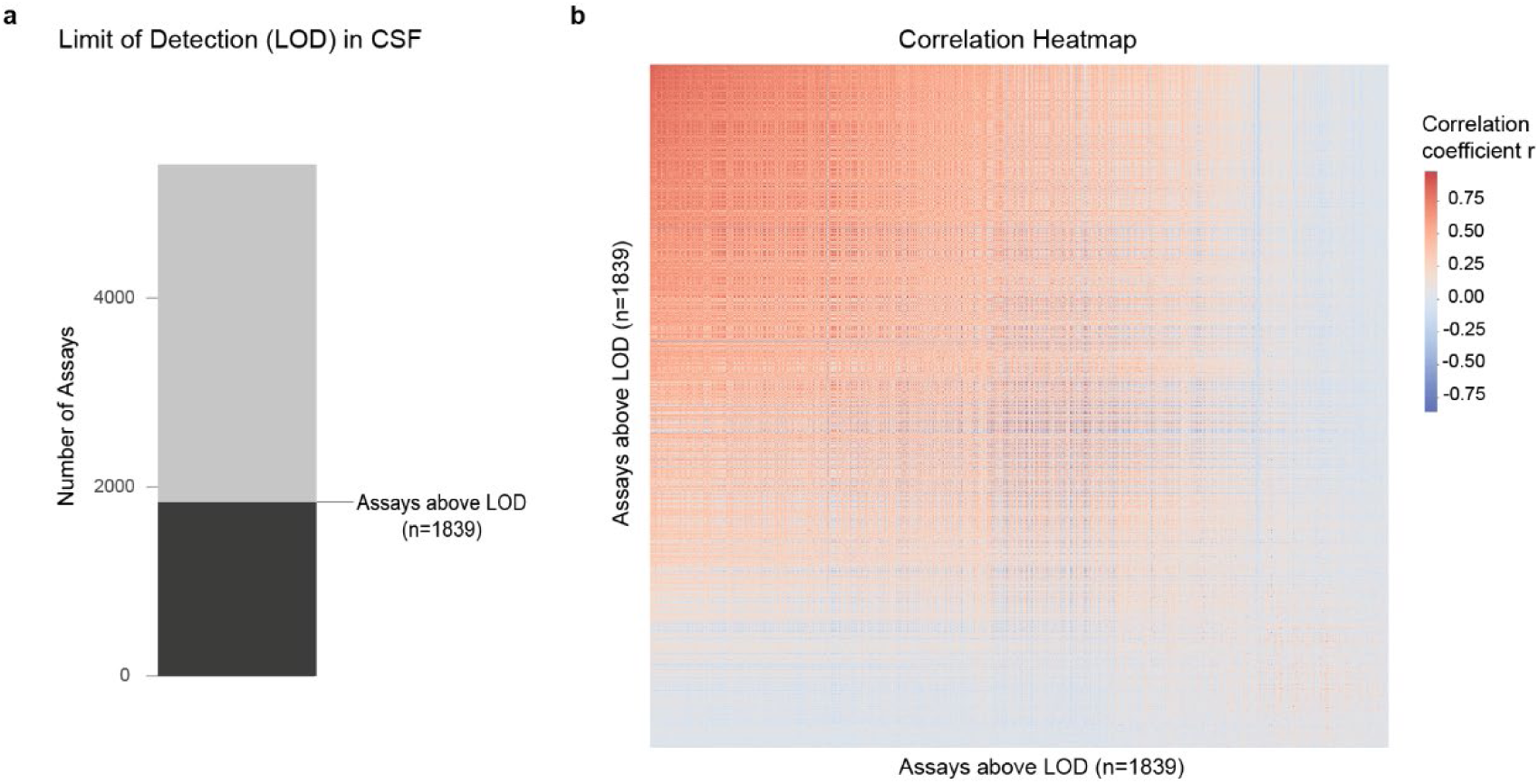
Empirical limit of detection (LOD) of the OlinkExplore HT assay in cerebrospinal fluid and correlation heatmap of the 1,839 measurable proteins. **a,** Stringent quality control measures of the proximity extension assay (PEA) OlinkExplore HT were implemented to ensure the reliability of the cerebrospinal fluid (CSF) proteomics dataset. Assays with NPX values below the limit of detection (LOD) in all samples were excluded (grey), resulting in a refined dataset of 1839 assays (black) for further analysis. **b,** Heatmap visualizing pairwise Pearson correlation coefficients (r) to illustrate the degree and direction of linear relationships among the NPX values of the 1839 proteins above LOD quality filtering.

**eFigure S3:**
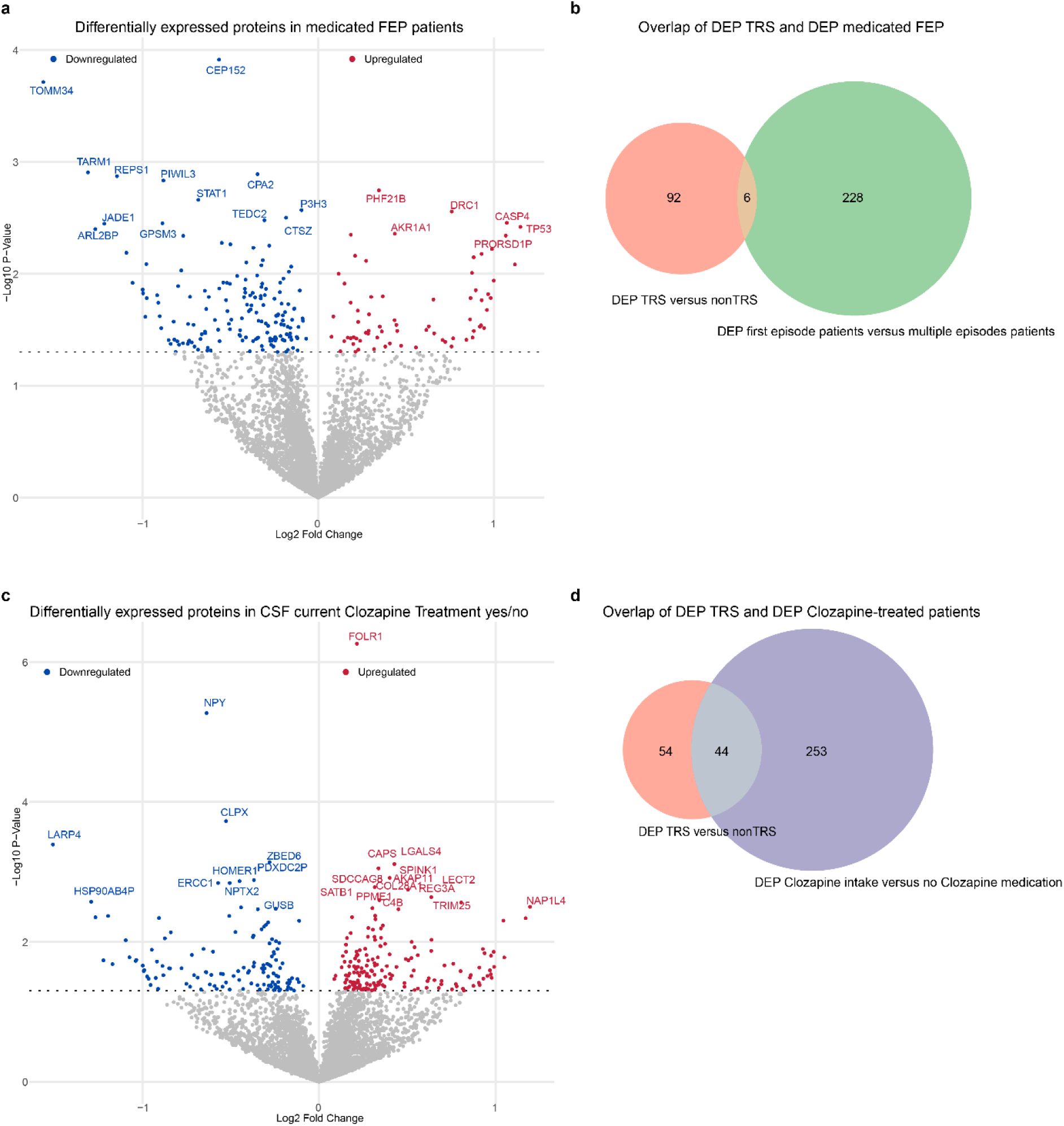
Proteomic subgroup profiling of Clozapine-treated and medicated first-episode patients reveals separate cluster of treatment-resistance-associated proteins. **a,** Volcano plot illustrating differential protein expression analysis using a two-sided linear model t-test (Limma) and comparing treated patients with first episode psychosis (FEP) to those with recurrent multiple episodes (MEP) who have experienced more than one psychotic episode, plotting the log2 fold change on the x-axis against the -log10 p-value on the y-axis. Dotted line indicates p-value threshold of 0.05. Differentially expressed proteins (DEP) with significant upregulation (red) and downregulation (blue) in FEPs exceeding a p-value threshold of 0.05 are color-coded, while non-significant proteins are shown in grey. **b,** Venn diagram visualizing the overlap of the TRS-DEP and DEP between FEP- and MEP-patients. Numbers represent N DEP. **c,** Volcano plot illustrating differential protein expression analysis using a two-sided linear model t-test (Limma) and comparing CSF of Clozapine-medicated compared to non-Clozapine-medicated patients, plotting the log2 fold change on the x-axis against the -log10 p-value on the y-axis. Dotted line indicates p-value threshold of 0.05. Proteins with significant upregulation (red) and downregulation (blue) are color-coded. **d,** Venn diagram visualizing the overlap of TRS-DEP and the DEP of Clozapine-medicated vs. non-Clozapine medicated patients. Numbers represent N DEP.

