## Supplement 1 for "Cerebrospinal fluid proteomics reveals synaptic and immune dysregulation in treatment-resistant schizophrenia"

*Fischer* et al.

Supplement 1 (this document):

- Legends supplementary tables: TableS1, TableS2, TableS3, TableS4, TableS5
    - separate Excel files
- eFigures 1–3
- eMethods

*Corresponding authors:*

*Laura E. Fischer, Max Planck Institute of Psychiatry, Kraepelinstr. 2-10, 80804 Munich, Germany, Mail:*

*Florian J. Raabe, Max Planck Institute of Psychiatry, Kraepelinstr. 2-10, 80804 Munich, Germany, Mail:*

### Supplementary tables

**Table S1. Baseline characteristics.** See separate excel file.

**Table S2. Lists of DEPs of Limma analysis and GSEA pathways.** See separate excel file.

**Table S3. Characterization of DEPs.** See separate excel file.

**Table S4. CLZ and CPZ correlation with NPX values.** See separate excel file.

**Table S5. Features of SVM.** See separate excel file.

### eFigures

*For submission attached to main manuscript*

### eMethods

#### Study design and participants

In center 1, patients were enrolled in a biobanking study between May 2001 and July 2023. Schizophrenia spectrum disorder (SSD) diagnosis was based on ICD-10, clinical characteristics and clinical trajectories were verified based on information provided in medical records screened independently by two trained psychiatrists. In center 2, patients were enrolled between August 2018 and April 2024. Inclusion criteria comprised age between 18 and 65 years and a diagnosis of SSD, according to DSM-V, assessed with the Mini International Neuropsychiatric Interview (M.I.N.I.), German version 7.0.2[1]. The study in center 1 was approved by the ethics committee of the Bavarian State Medical Association (Bayerische Landesaerztekammer) (vote 2010-069). The study in center 2 was approved by the local ethics committee of the Ludwig Maximilian University Munich (vote 17-061, 21-0183, 21-1139 and vote 18-716[2]). The entire study was conducted according to the Declaration of Helsinki. Written informed consent was obtained from all participants before taking part in the study. Mainly patients with SCZ (N=136) and some patients with schizoaffective disorder (N=6) and brief psychotic disorder (N=6) were recruited. We retrospectively classified all patients into the TRS and nonTRS groups (**eFigure S1A**) guided by the TRRIP (Treatment Response and Resistance in Psychosis) consensus criteria[3] and in line with recommendations for real-world data[4] as following: In both centers, with regard to antipsychotic dosage, an equivalent dose ≥ 600mg of chlorpromazine equivalents (≈18 mg olanzapine) per day-was considered as a minimum requirement for treatment resistance based on established conversion criteria[5-9]).

Regarding treatment adherence, all patients were hospitalized under standard monitoring protocols at both centers, which included routine therapeutic drug monitoring and daily oversight by hospital staff to help maintain regular medication intake.

In Center 1, treatment duration could not be directly assessed, so we defined TRS as the use of two antipsychotic medications at adequate doses concurrently. The criterion of at least moderate severity of illness was met by all patients, as they were all hospitalized due to their symptoms. In Center 1, Non-Remission was evaluated on the basis of the PANSS Remission in Schizophrenia Working Group (RSWG) items without the time criterion ("Andreasen criteria")[10]. In Center 2, "adequate pharmacological treatment" was defined as antipsychotic therapy for ≥8 weeks, including at least four weeks with two different agents either concurrently or sequentially. Non-remission was determined by two trained psychiatrists through a comprehensive review of patient records where it was assessed by the psychiatrists involved in the patient's care. Additionally, receiving CLZ treatment at any point in their lifetime or currently was also used as a defining criterion for TRS. For more information regarding the screening algorithms of clinical records, please see **eFigure S1b**. In both centers, first-episode psychosis patients were identified as those who underwent lumbar puncture during their initial psychotic episode. Multi-episode patients were those who had experienced more than one psychotic episode at the timepoint of the lumbar puncture. As the long-term aim of our group is to develop clinically applicable biomarker signatures to predict patients on risk for TRS, we only excluded patients with concurrent neurological disease at the timepoint of LP.

#### CSF collection and pre-analytic handling

CSF assembly was embedded in diagnostic lumbar punctures within the clinical routine care, following the national schizophrenia guideline[11,12]. Cerebrospinal fluid (CSF) was obtained via lumbar puncture at either the L4–L5 or L5–S1 interspace. We excluded CSF samples with erythrocyte counts above 500/µL in order to avoid artificial carry-over effects of serum proteins into the CSF. For center 1: CSF samples (Polypropylene Tubes®, 2 mL) were processed within 2 hours. The samples were centrifuged at 200 × g for 20 minutes at 8°C to pellet and remove cells. The supernatant was stored at -80°C immediately after processing for further analyses. For center 2: The CSF samples (Polypropylene Tubes®, 2 mL) were processed within one hour. The samples were centrifuged at 400 × g for 10 minutes at 4°C to pellet and remove cells, leaving the supernatant for subsequent analysis. The supernatant was immediately frozen and stored at -80°C for further analyses.

For the proteomic investigation, all samples were thawed on ice and transferred to 96-well plates for further proteomic analysis.

#### Proteomics data pre-processing

For the proteomics data pre-processing, the mean expression value was calculated for proteins that were measured in multiple blocks. Only measurements that passed both assay and sample quality control were included in the analysis. Only proteins that exceeded the limit of detection (LOD, normalized manufacturer-provided fixed LOD values for each assay) in at least one sample, as determined by the *OlinkAnalyze R package (v* 4.2.0), were included in the analysis. For proteins with partially missing values (in some samples below the limit of detection (LOD), the NPX values were retained to preserve quantitative information and minimize data distortion.

#### Statistical analysis

Significant differences between the two cohorts for the characteristics presented in **Table 1** and **Table S1** were assessed using two-sided Fisher's exact test (fisher_exact from the python package SciPy 1.13.0) for contingency tables or a two-sided Mann–Whitney U test (mannwhitneyu from the python package SciPy 1.13.0) for continuous values.

#### Correlation Analysis of Protein Expression

To assess the co-regulation patterns among the measured proteins, we calculated pairwise Pearson correlation coefficients across all proteins (function *pandas.DataFrame.corr()* from the python package pandas 2.2.3). To focus solely on inter-protein relationships and exclude trivial self-correlations, the diagonal elements of the resulting correlation matrix were set to NaN.

#### Differential expression analysis

When applying the *limma R package (v 3.58.1)*[13,14], a linear model was applied to the data, followed by the computation of empirical Bayes moderated t-statistics adjusted for sex, age, and center. A significance threshold of $p < 0.05$ was set to identify differentially expressed proteins (DEPs). Subsequently, we used the *simpleM* approach from Gao et al. 2008 which corrects for non-independence among correlated features while maintaining control of type I error rates[15], using the respective *R package (v poolr_1.1-1)*, which calculates the effective number (n(eff)) of independent tests based on the eigenvalues of a correlation matrix[15]. By employing spectral decomposition through principal component analysis (PCA), we reduced the dimensionality of the data and determined n(eff) as the number of principal components explaining 99.5% of the total variance. This adjustment allowed for a more accurate calculation of the significance threshold by dividing the nominal alpha level by n(eff), effectively controlling the experiment-wide error rate. Log2 fold-change (log2FC) values for all proteins were computed as part of the differential expression analysis to quantify the magnitude of changes in protein expression.

Characterization of the DEPs was performed using the Open Targets Platform[16]. Links were uncovered via a Europe PMC text-mining pipeline using an evidence-scoring scheme that weights document sections, sentence contexts, and titles, with each gene–disease co-occurrence score normalized to the 0–1 range. Additional protein annotations were obtained from Olink Proteomics. (2025). Olink Insight Data Analysis Platform, Uppsala, Sweden. Retrieved from https://www.olink.com/products/insight/ (access 31.01.2025), which provides supplementary information on protein function and disease associations.

#### Gene set enrichment analysis of DEPs

Gene Set Enrichment Analysis (GSEA) with the Gene ontologies "Biological processes", "Molecular function", and "Cellular components"[17,18] was performed using the *OlinkAnalyze R package (v* 4.2.0, olink_pathway_enrichment function, integrating clusterProfiler[19-21] for pathway analysis[22]. Protein identifiers and statistical test results (fold changes and adjusted p-values from the differential expression analysis) of all assays measured (background/context) were considered for GSEA. Pathway enrichment was assessed using Gene Ontology (GO) gene sets from Molecular Signatures Database (MSigDB) v2024.1.Hs (date of download 15.01.2025) [23] focusing on biological process (BP) and cellular compartment (CC) annotation. All genes within the top specificity decile and all GO terms with a gene set size of 10 to 500 genes were used as input. The GSEA method was applied to rank proteins based on their normalized enrichment score (NES). Enrichment was considered significant for pathways with a NES greater than 1 or less than -1 and a q-value < 0.25[24].

The identified DEPs were additionally cross-referenced with Synaptic Gene Ontologies (SynGO)[25] (release 1.2, '20231201') synaptic gene sets to evaluate their functional associations with specific synaptic compartments and biological processes[25].

#### Multi-marker Analysis of GenoMic Annotation (MAGMA) analysis

MAGMA (Multi-marker Analysis of GenoMic Annotation, version 1.09)[26] was used to carry out formal enrichment analyses using the summary statistics of schizophrenia[27] and treatment-resistant schizophrenia GWAS [28]. DEPs in our Limma analyses defined our gene-sets of interest. After SNP-to-gene annotation, gene-level analyses (±10kb) were carried out using the "SNPwise-mean" model. Gene-based p-values were obtained by combining the SNP p-values in a gene (±10kb) into a gene test statistic (mean $\chi 2$) corrected for linkage disequilibrium between SNPs, gene size, and density. MAGMA competitive gene-set analysis tested, within a regression framework, if the DEPs gene-sets have a stronger association with the target phenotype than randomly selected gene-sets with similar characteristics. For gene-set competitive testing, linear regression analyses were conditioned to gene size, log (gene size), gene density, log (gene density), inverse mac and log (inverse mac).

#### Support Vector Machine (SVM) model for predicting treatment resistance

To explore the patient-level predictability of TRS based on CSF proteomics data above the LOD threshold, we employed the machine learning tool NeuroMiner (https://github.com/neurominer-git/NeuroMiner_1.3). First, we applied partial correlation to adjust for potential confounding factors, including age and sex. This step ensured that downstream analyses focused on the variance most relevant to treatment resistance. Next, we scaled all feature values to the 0-1 range. We then ranked features by their f-score relative to the target label, defined as *f-score = (mean (TRS) – mean (Non-TRS) / (sd (TRS)+sd (Non-TRS))*. Based on these ranks, we applied thresholding at various percentile cutoffs (e.g., 80%, 75%, 95%) to reduce the feature space to the most discriminative proteins. Following this, we performed median-based standardization and applied winsorization at ±4 standard deviations to mitigate the impact of extreme outliers. For classification, we employed an L2-regularized L1-loss Support Vector Classifier (SVC) in its dual formulation, implemented via LIBLINEAR with a convergence tolerance of 0.01. This method balances a robust penalty on misclassifications (L1 loss) with coefficient shrinkage (L2 regularization), enhancing generalization performance.

We conjointly optimized the hyperparameters of the preprocessing (percentile thresholds) and model training (*C* parameter) steps using grid-search. Preprocessing optimization involved four parameter combinations (e.g., threshold cutoffs), while model optimization tested 21 different *C* parameters ($C \in 2^{[-13 \rightarrow 0]}$). A repeated nested cross-validation scheme (CV2: 10×10 outer folds; CV1: 5×10 inner folds) was used to reliably assess generalization performance, minimizing overfitting and ensuring robust estimation of classification metrics.

Performance metrics, including accuracy, sensitivity, specificity, balanced accuracy (BAC), area under the curve (AUC), and Matthews Correlation Coefficient (MCC), were calculated to evaluate the performance of the model. Confusion matrices were generated to assess classification results, including true positive, true negative, false positive, and false negative rates. Statistical measures such as Youden's J, Proportional Success Index, and likelihood ratios (+LR, -LR) were derived to quantify classification robustness.

Permutation analysis was applied to test statistical significance of the observed classification performance as described by Koutsouleris et al., 2016[29]. Random permutations of the outcome label were performed 1000 times and the SVM models were retrained for each permutation using the established CV-scheme. By accumulating the predictions from the random models into a permuted ensemble prediction for each CV2 subject, a null distribution of classification performance was constructed for the prediction model. Significance of the observed model performance (BAC) was calculated by dividing the number of events where the permuted BAC was higher or equal to the observed BAC by the number of permutations performed. Model significance was determined at $\alpha=0.05$.

#### References Supplement 1

7. Leucht S, Samara M, Heres S, et al. Dose Equivalents for Second-Generation Antipsychotic Drugs: The Classical Mean Dose Method. *Schizophr Bull*. Nov 2015;41(6):1397-402. doi:10.1093/schbul/sbv037
8. Leucht S, Samara M, Heres S, Davis JM. Dose Equivalents for Antipsychotic Drugs: The DDD Method. *Schizophr Bull*. Jul 2016;42 Suppl 1(Suppl 1):S90-4. doi:10.1093/schbul/sbv167
9. Leucht S, Crippa A, Siafis S, Patel MX, Orsini N, Davis JM. Dose-Response Meta-Analysis of Antipsychotic Drugs for Acute Schizophrenia. *Am J Psychiatry*. Apr 1 2020;177(4):342-353. doi:10.1176/appi.ajp.2019.19010034
10. Andreasen NC, Carpenter WT, Jr., Kane JM, Lasser RA, Marder SR, Weinberger DR. Remission in schizophrenia: proposed criteria and rationale for consensus. *Am J Psychiatry*. Mar 2005;162(3):441-9. doi:10.1176/appi.ajp.162.3.441
11. Gaebel W, Falkai P, Hasan A. The revised German evidence- and consensus-based schizophrenia guideline. *World Psychiatry*. Feb 2020;19(1):117-119. doi:10.1002/wps.20706
12. Gaebel W, Hasan A, Falkai P. *S3-Leitlinie Schizophrenie*. Springer Berlin Heidelberg; 2019.
13. Phipson B, Lee S, Majewski IJ, Alexander WS, Smyth GK. ROBUST HYPERPARAMETER ESTIMATION PROTECTS AGAINST HYPERVARIABLE GENES AND IMPROVES POWER TO DETECT DIFFERENTIAL EXPRESSION. *Ann Appl Stat*. Jun 2016;10(2):946-963. doi:10.1214/16-aoas920
14. Ritchie ME, Phipson B, Wu D, et al. limma powers differential expression analyses for RNA-sequencing and microarray studies. *Nucleic Acids Res*. Apr 20 2015;43(7):e47. doi:10.1093/nar/gkv007
15. Gao X, Starmer J, Martin ER. A multiple testing correction method for genetic association studies using correlated single nucleotide polymorphisms. *Genet Epidemiol*. May 2008;32(4):361-9. doi:10.1002/gepi.20310
16. Buniello A, Suveges D, Cruz-Castillo C, et al. Open Targets Platform: facilitating therapeutic hypotheses building in drug discovery. *Nucleic Acids Res*. Jan 6 2025;53(D1):D1467-d1475. doi:10.1093/nar/gkae1128
17. Ashburner M, Ball CA, Blake JA, et al. Gene ontology: tool for the unification of biology. The Gene Ontology Consortium. *Nat Genet*. May 2000;25(1):25-9. doi:10.1038/75556
18. Aleksander SA, Balhoff J, Carbon S, et al. The Gene Ontology knowledgebase in 2023. *Genetics*. May 4 2023;224(1)doi:10.1093/genetics/iyad031
19. Yu G. Thirteen years of clusterProfiler. *The Innovation*. 2024;5(6)doi:10.1016/j.xinn.2024.100722
20. Yu G, Wang LG, Han Y, He QY. clusterProfiler: an R package for comparing biological themes among gene clusters. *Omics*. May 2012;16(5):284-7. doi:10.1089/omi.2011.0118
21. Wu T, Hu E, Xu S, et al. clusterProfiler 4.0: A universal enrichment tool for interpreting omics data. *Innovation (Camb)*. Aug 28 2021;2(3):100141. doi:10.1016/j.xinn.2021.100141
22. Korotkevich G, Sukhov V, Sergushichev A. Fast gene set enrichment analysis. *bioRxiv*. 2019:060012. doi:10.1101/060012
23. Liberzon A, Birger C, Thorvaldsdóttir H, Ghandi M, Mesirov JP, Tamayo P. The Molecular Signatures Database (MSigDB) hallmark gene set collection. *Cell Syst*. Dec 23 2015;1(6):417-425. doi:10.1016/j.cels.2015.12.004
24. Subramanian A, Tamayo P, Mootha VK, et al. Gene set enrichment analysis: a knowledge-based approach for interpreting genome-wide expression profiles. *Proc Natl Acad Sci U S A*. Oct 25 2005;102(43):15545-50. doi:10.1073/pnas.0506580102
25. Koopmans F, van Nierop P, Andres-Alonso M, et al. SynGO: An Evidence-Based, Expert-Curated Knowledge Base for the Synapse. *Neuron*. Jul 17 2019;103(2):217-234.e4. doi:10.1016/j.neuron.2019.05.002
26. de Leeuw CA, Mooij JM, Heskes T, Posthuma D. MAGMA: generalized gene-set analysis of GWAS data. *PLoS Comput Biol*. Apr 2015;11(4):e1004219. doi:10.1371/journal.pcbi.1004219

27. Trubetskoy V, Pardiñas AF, Qi T, et al. Mapping genomic loci implicates genes and synaptic biology in schizophrenia. *Nature*. Apr 2022;604(7906):502-508. doi:10.1038/s41586-022-04434-5
28. Pardiñas AF, Smart SE, Willcocks IR, et al. Interaction Testing and Polygenic Risk Scoring to Estimate the Association of Common Genetic Variants With Treatment Resistance in Schizophrenia. *JAMA Psychiatry*. 2022;79(3):260-269. doi:10.1001/jamapsychiatry.2021.3799
29. Koutsouleris N, Kahn RS, Chekroud AM, et al. Multisite prediction of 4-week and 52-week treatment outcomes in patients with first-episode psychosis: a machine learning approach. *Lancet Psychiatry*. Oct 2016;3(10):935-946. doi:10.1016/s2215-0366(16)30171-7
